# Increased cardiovascular mortality in patients with mechanically expandable transcatheter aortic valve and without permanent pacemaker

**DOI:** 10.1101/2023.06.10.23291226

**Authors:** Petr Hájek, Radka Adlová, Martin Horváth, Eva Hansvenclová, Monika Pecková, Josef Veselka

## Abstract

**Introduction:** Use of the mechanically expandable transcatheter aortic valve (MEV) has been recently linked to increased risks of valve dysfunction and cardiovascular mortality. The risk of developing conduction disturbance with the MEV valve is well-known, and the negative prognostic impact of permanent pacemaker implantation (PPI) after transcatheter aortic valve implantation (TAVI) is another consideration.

**Aim:** This study aimed to compare the mid-term survival of patients with MEV and self-expandable valves (SEV), and to examine survival of both groups according to the presence or absence of PPI.

**Methods:** This single-center, retrospective, observational study examined data from MEV and SEV groups comprising 92 and 373 patients, respectively. The mean clinical follow-up was 2.5 ± 1.7 years. Mortality information was obtained from the National Institutes of Health Information and Statistics.

**Results:** Baseline characteristics were comparable between the groups. The log-rank test showed higher cardiovascular mortality in the MEV group [*p*=0.042; RR: 1.594 (95%CI: 1.013-2.508)]. The Cox proportional hazards model identified MEV implantation as an independent predictor of cardiovascular mortality. The rate of PPI was twice as high in the MEV versus SEV group (33.7% vs. 16.1%; *p* <0.001). We compared the survival of both groups according to the presence or absence of PPI and found higher mortality in the MEV group without PPI versus the SEV group without PPI (*p*=0.007; RR: 2.156 [95%CI: 1.213-3.831]). Survival did not differ in the groups with PPI.

**Conclusions:** A higher mid-term cardiovascular mortality rate was observed with MEV versus SEV implants. Comparing both groups according to the presence or absence of PPI, we observed a higher mortality risk in patients with MEV without PPI than in SEV without PPI. In contrast, mortality did not differ between the groups when PPI was implanted.

**What is already known on this topic:** Use of the mechanically expandable transcatheter aortic valve (MEV) has been recently linked to increased risks of valve dysfunction and cardiovascular mortality. The risk of developing conduction disturbance with the MEV valve is well-known, and the negative prognostic impact of permanent pacemaker implantation (PPI) after transcatheter aortic valve implantation is another consideration.

**What this study adds:** Our study suggested that higher cardiovascular mortality was independently associated with the use of MEV versus self-expandable valves (SEV) implants. Comparing both groups according to the presence or absence of PPI, we observed a higher mortality risk in patients with MEV without PPI than in those with SEV without PPI.

**How this study might affect research, practice or policy:** Although MEV were recalled in 2020, thousands of patients have been treated with them. Therefore, patients with MEV without PPI deserve increased attention during long-term follow-up.

## Introduction

Recently published data have revealed a higher risk of bioprosthetic valve dysfunction of mechanically expandable valves (MEV) designed for transcatheter aortic valve implantation (TAVI) (1, 2). In fact, the results of our pilot study (3) have suggested that higher cardiovascular mortality was independently associated with the use of MEV versus self-expandable transcatheter valves (SEV) over mid-term follow-up. Although MEV were recalled in 2020 (4,5) and are not currently available for clinical use, more than 10000 patients have been treated with them (6). To date, long-term MEV data have been published from only one randomized clinical trial (6). The authors confirmed a higher risk of prosthetic thrombosis with MEV implants, but they found comparable survival outcomes to those with SEV.

New permanent pacemaker implantation (PPI) has been studied as a potential risk factor for long-term survival after TAVI (7). Also, MEV implantation has been associated with a high risk of conduction disturbances (8, 9). Because patients with MEV have constituted only a small part of the published meta-analyses (7), it is unclear whether the results regarding the effect of PPI on survival after TAVI are applicable.

### Aim

The aim of our study was to compare the mid-term survival of patients with MEV and SEV and to evaluate survival of both groups according to the presence or absence of PPI.

## Methods

### Design

We conducted a single-center, retrospective, observational study comparing the outcomes of patients with severe aortic stenosis (AS) who underwent TAVI with mechanically expandable intra-annular Lotus® and Lotus Edge® (Boston Scientific, USA) versus self-expandable supra-annular Evolut R® (Medtronic, U.S.A.) and Acurate Neo® (Boston Scientific, USA) transcatheter valves. The study was approved by a multicenter ethics committee and conducted in accordance with the principles of the Declaration of Helsinki. All participants signed informed consent.

### Patients and procedures

A total of 481 consecutive patients with severe AS who underwent TAVI between August 2015 and June 2022 were enrolled. On admission, patients were either hemodynamically stable for planned diagnostic evaluation or acutely decompensated with heart failure. Severe AS was diagnosed according to echocardiographic criteria. The indication for TAVI was established by consensus among members of the Heart Team. All TAVI procedures were performed by highly experienced interventional cardiologists at a single tertiary center. Clinical, demographic, and echocardiographic data were recorded at baseline and over follow-up. Serum troponin I levels were routinely analyzed 24 h after TAVI. The patients underwent post-TAVI clinical and echocardiographic examinations prior to hospital discharge, at 30 days, and at one year. Information regarding specific events was obtained from follow-up visits and the National Institutes of Health Information and Statistics.

The Lotus® and Lotus Edge® implants are bioprosthetic aortic valves comprising a braided nitinol wire frame with three bovine pericardial leaflets and a polymer membrane that surrounds the lower valve half to reduce paravalvular leaks. The valve is pre-mounted on a delivery catheter and deployed via controlled mechanical expansion, enabling repositioning or retrieval of the valve at any point before its release (10). The novel features of the Lotus Edge® system include increased flexibility of the delivery catheter, enhanced visualization of the locking mechanism, and Depth Guard technology designed to reduce left ventricular outflow tract (LVOT) interactions and potentially reduce PPI (11).

The Medtronic CoreValve Evolut R®System comprises the Evolut R valve and the EnVeo R Delivery Catheter System (DCS) with the InLine sheath. The trileaflet valve and sealing skirt are made of porcine pericardial tissue and sutured in a supra-annular position on a compressible and self-expandable nitinol frame. EnVeo R DCS enables the valve to be fully repositionable and recapturable before full release by turning the delivery handle (12). The Acurate Neo® (Boston Scientific, USA) transcatheter valve consists of three porcine pericardial leaflets mounted on a self-expanding nitinol frame with an upper crown that provides supra-annular anchoring and caps the native leaflets, a waist that conforms to the native annulus, and a lower crown protruding few millimeters into the LVOT; an inner and outer porcine pericardium fabric skirt covers the inflow tract of the nitinol stent (13). All TAVI procedures were performed using the Lotus® Introducer set (Boston Scientific).

### Outcomes

The primary outcomes were the mid-term all-cause and cardiovascular mortality rates. Secondary outcomes were clinical outcomes including periprocedural myocardial infarction; overt central nervous system injury at 30 days; bleeding complications type 2, 3, or 4 at 30 days; major vascular complications at 30 days; acute kidney failure stage 2 or 3; and valve malposition. Outcomes were defined according to standardized endpoint definitions for transcatheter aortic valve implantation clinical trials in a consensus report from the Valve Academic Research Consortium (VARC3) (14).

### Statistical analysis

Data are presented as mean ± standard deviation (SD) for continuous variables and as median ± interquartile range for continuous variables with outliers. Continuous variables were compared using Welch’s two-sample t-tests for normally distributed variables and two-sample Wilcoxon tests for variables with some outliers. Categorical variables were compared using the Pearson chi-square test of independence or Fisher’s exact test (in case of small, expected counts). *P*-value<0.05 was considered statistically significant.

The log-rank test was used to compare survival times between the two groups. Kaplan-Meier estimates of the survival curves are presented. The Cox proportional hazards model was used to identify predictors of all-cause and cardiovascular mortality and to adjust for potential baseline differences between the groups. We analyzed data including sex, age, presence of diabetes mellitus, arterial hypertension, atrial fibrillation, coronary artery disease, creatinine level before and after TAVI, hemoglobin and platelet levels before and after TAVI, Society of Thoracic Surgeons (STS) score, aortic valve gradient (AVG) before and after TAVI, ejection fraction, aortic valve area before TAVI, wall thickness, NYHA classification before TAVI, necessity of acute TAVI, aortic regurgitation before TAVI, balloon valvuloplasty, amount of contrast dye during TAVI, open surgical access, pacemaker before and after TAVI, and type of TAVI valve. All analyses were performed using the statistical program R and GraphPad Prism Version 6.05.

## Results

### Baseline characteristics

A total of 481 consecutive single-center patients with symptomatic AS received the Lotus®, Lotus Edge®, Evolut R®, or Acurate Neo® valves. Valve selection was made at the discretion of clinicians. Sixteen patients were excluded (3.3%; 12 patients with valve-in-valve procedure, four with pure aortic regurgitation). Therefore, our sample comprised 465 patients (72 Lotus® and 20 Lotus Edge® valves in the MEV group; and 167 Evolut R® and 206 Acurate Neo® valves in the SEV group). The baseline clinical and echocardiographic characteristics did not differ between groups, except for a slightly higher degree of left ventricle hypertrophy present in the MEV group (**Table 1**).

**Table 1.** Clinical and echocardiographic characteristics.

|  | SEV group<br>All<br>N = 373 | MEV group<br>All<br>N = 92 | *p*-value | SEV group<br>PM +<br>N = 103 | MEV group<br>PM +<br>N = 40 | *p*-value | SEV group<br>PM -<br>N = 270 | MEV group<br>PM -<br>N = 52 | *p*-value |
| --- | --- | --- | --- | --- | --- | --- | --- | --- | --- |
| **Clinical characteristics** |  |  |  |  |  |  |  |  |  |
| Age, years * | 77.5 ± 7.3 | 78.8 ± 6.4 | 0.113 | 79.4 ± 6.4 | 79.0 ± 7.0 | 0.767 | 76.8 ± 7.5 | 78.6 ± 6.0 | 0.072 |
| Males, n (%) | 199 (53.4) | 48 (52.2) | 0.839 | 68 (66) | 28 (70) | 0.696 | 131 (48.5) | 20 (38.5) | 0.183 |
| Body mass index * | 29.0 ± 5.3 | 28.7 ± 4.7 | 0.610 | 28.4 ± 5.1 | 28.9 ± 5.0 | 0.541 | 29.2 ± 5.4 | 28.5 ± 4.5 | 0.314 |
| NYHA class † | 2.5 ± 1.0 | 3.0 ± 1.0 | 0.538 | 2.5 ± 1.0 | 3.0 ± 1.0 | 0.934 | 2.5 ± 1.0 | 3.0 ± 1.0 | 0.532 |
| NYHA class ≥ III, n (%) | 165 (45.2)<sup>#</sup> | 50 (54.9)<sup>#</sup> | 0.096 | 47 (47.0)<sup>#</sup> | 21 (53.8)<sup>#</sup> | 0.468 | 118 (44.5)<sup>#</sup> | 29 (55.8) | 0.137 |
| Diabetes mellitus, n (%) | 156 (41.9) | 30 (32.6) | 0.102 | 50 (48.5) | 12 (30.0) | 0.045 | 106 (39.4) | 18 (34.6) | 0.516 |
| Arterial hypertension, n (%) | 301 (80.7) | 68 (73.9) | 0.136 | 90 (87.4) | 30 (75.0) | 0.071 | 211 (78.4) | 38 (73.1) | 0.396 |
| Atrial fibrillation, n (%) | 126 (33.8) | 35 (38.0) | 0.441 | 55 (53.4) | 21 (52.5) | 0.923 | 71 (26.3) | 14 (26.9) | 0.925 |
| Coronary artery disease, n (%) | 172 (46.1) | 37 (40.2) | 0.308 | 49 (47.6) | 20 (50.0) | 0.794 | 123 (45.6) | 17 (32.7) | 0.087 |
| Creatinine, µmol/l † | 88 ± 37 | 87 ± 42 | 0.951 | 95 ± 51 | 89 ± 61 | 0.332 | 84 ± 31 | 87 ± 37 | 0.995 |
| Hemoglobin, g/l * | 125.5 ± 17.6 | 125.9 ± 16.1 | 0.814 | 125.4 ± 17.8 | 126.7 ± 17.4 | 0.704 | 125.5 ± 17.6 | 125.4 ± 15.2 | 0.950 |
| Platelets, x10<sup>9</sup>/l * | 207 ± 67 | 192 ± 66 | 0.067 | 191 ± 71 | 180 ± 60 | 0.404 | 213 ± 64 | 202 ± 69 | 0.269 |
| Pacemaker before TAVI, n (%) | 43 (11.5) | 9 (9.8) | 0.715 | 43 (41.7) | 9 (22.5) | 0.032 | - | - | - |
| LBBB before TAVI, n (%) | 13 (3.5) | 7 (7.6) | 0.081 | 0 | 1 (2.5) | 0.107 | 13 (4.8) | 6 (11.5) | 0.060 |
| STS score † | 2.4 ± 2.7 | 2.2 ± 2.3 | 0.350 | 2.7 ± 3.3 | 2.3 ± 2.4 | 0.140 | 2.2 ± 2.5 | 2.1 ± 2.3 | 0.583 |
| Acute procedure, n (%) | 38 (10.2) | 7 (7.6) | 0.454 | 9 (8.7) | 2 (5.0) | 0.452 | 29 (10.7) | 5 (9.6) | 0.809 |
| Agatston score † | 2441 ± 1796 | 3213 ± 2431 | 0.101 | 2637 ± 1963 | 3316 ± 2014 | 0.327 | 2382 ± 1772 | 3088 ± 2947 | 0.264 |
| **Echocardiography characteristics** |  |  |  |  |  |  |  |  |  |
| Mean gradient, mmHg * | 42 ± 14 | 43 ± 15 | 0.298 | 40 ± 14 | 42 ± 14 | 0.679 | 42 ± 14 | 45 ± 15 | 0.235 |
| Aortic valve area, cm<sup>2</sup> * | 0.8 ± 0.2 | 0.7 ± 0.2 | 0.175 | 0.8 ± 0.2 | 0.8 ± 0.2 | 0.480 | 0.8 ± 0.2 | 0.7 ± 0.2 | 0.166 |
| Aortic regurgitation grade ≥ 3, n (%) | 16 (4.4)<sup>#</sup> | 5 (5.4) | 0.587 | 3 (3.0)<sup>#</sup> | 1 (2.5) | 1.000 | 13 (4.9)<sup>#</sup> | 4 (7.7) | 0.495 |
| LVEF, % † | 60 ± 15 | 60 ± 20 | 0.365 | 55 ± 25 | 55 ± 15 | 0.500 | 60 ± 15 | 60 ± 15 | 0.258 |
| Septal wall thickness, mm * | 12.6 ± 2.1 | 13.2 ± 2.0 | 0.014 | 12.8 ± 2.1 | 13.3 ± 2.2 | 0.195 | 12.5 ± 2.1 | 13.1 ± 1.9 | 0.048 |
| Posterior wall thickness, mm * | 11.8 ± 1.6 | 12.4 ± 1.5 | <0.001 | 11.9 ± 1.6 | 12.4 ± 1.5 | 0.096 | 11.8 ± 1.6 | 12.5 ± 1.4 | 0.002 |
| Mitral regurgitation grade ≥ 3, n (%) | 39 (10.6)<sup>#</sup> | 7 (7.6) | 0.397 | 19 (18.6)<sup>#</sup> | 4 (10.0) | 0.209 | 20 (7.5)<sup>#</sup> | 3 (5.8) | 0.660 |

\* Plus-minus values are mean ± SD; † Plus-minus values are median ± interquartile range; <sup>#</sup> Percentage is calculated from the available number of examinations performed. LBBB, left bundle branch block; LVEF = *Left ventricular ejection fraction*; MEV, mechanically expandable valve; NYHA class, New York Heart Association classification; PM +, permanent pacemaker implanted; PM −, permanent pacemaker not implanted; SEV, self-expandable valve; STS, Society of Thoracic Surgeons score.

### Procedural and post-procedural characteristics (Table 2)

In the SEV group, we used balloon valvuloplasty more often and a larger amount of contrast dye. Open surgical access was used more often in the MEV group. We observed a slight but significantly greater drop in hemoglobin and platelet counts in the MEV group. The mean AVG (AVGm) at 30 days was significantly higher in the MEV group, and this difference remained significant at one year.

In the MEV group, there were two-fold rates of new PPI, new onset of left bundle branch block (NO-LBBB), and increased cardiac troponin I levels compared to the SEV group. In the SEV group, higher troponin I levels were observed in those with new PPI versus those without (430 ± 1034 ng/l vs. 266 ± 542 ng/l, respectively ; *p*=0.001). In the MEV group, this difference was not statistically significant (648 ± 532 ng/l vs. 605 ± 1423 ng/l, respectively; *p*=0.916).

### Secondary outcomes

The rate of procedural complications was low and did not differ between the groups (Table 3). In addition, we observed the same technical success in the MEV and SEV groups (96.7% vs. 97.6%, respectively; *p*=0.712). The device success was numerically higher in the SEV group (92.2% SEV vs. 85.9% MEV; *p*=0.057). This difference was the result of the higher incidence of AVGm> 20 mmHg in the MEV group (Table 2). Significantly higher early safety composite endpoints were observed in the SEV group (77.4% SEV vs. 57.6% MEV; *p*<0.001). This difference was mainly driven by higher rates of PPI in the MEV group (33.7% MEV vs. 16.1% SEV; *p*<0.001) (Table 2). All-cause and cardiovascular mortality at 30 days and over the first year did not significantly differ (Table 3).

**Table 2.** Procedural and postprocedural characteristics.

|  | SEV group<br>All<br>N = 373 | MEV group<br>All<br>N = 92 | *p*-value | SEV group<br>PM +<br>N = 103 | MEV group<br>PM +<br>N = 40 | *p*-value | SEV group<br>PM -<br>N = 270 | MEV group<br>PM -<br>N = 52 | *p*-value |
| --- | --- | --- | --- | --- | --- | --- | --- | --- | --- |
| **Procedural characteristics** |  |  |  |  |  |  |  |  |  |
| Balloon valvuloplasty, n (%) | 180 (48.3) | 10 (11.1) | <0.001 | 47 (45.6) | 5 (12.5) | <0.001 | 133 (49.3) | 5 (10.0) | <0.001 |
| Open surgical access, n (%) | 118 (31.6) | 72 (78.3) | <0.001 | 42 (40.8) | 28 (70.0) | 0.002 | 76 (28.1) | 44 (84.6) | <0.001 |
| Contrast dye, ml † | 130 ± 60 | 100 ± 55 | <0.001 | 135 ± 70 | 110 ± 77 | 0.195 | 130 ± 60 | 100 ± 50 | <0.001 |
| **Postprocedural characteristics** |  |  |  |  |  |  |  |  |  |
| Troponin in 24 hours, ng/l † | 333 ± 593 | 634 ± 769 | <0.001 | 430 ± 1034 | 648 ± 532 | 0.076 | 266 ± 542 | 605 ± 1423 | <0.001 |
| Creatinine, µmol/l † | 77 ± 39 | 83 ± 41 | 0.257 | 92 ± 55 | 88 ± 48 | 0.732 | 75 ± 34 | 79 ± 37 | 0.320 |
| Hemoglobin, g/l * | 115 ± 18 | 111 ± 19 | 0.056 | 114 ± 18 | 111 ± 17 | 0.236 | 115 ± 18 | 111 ± 21 | 0.161 |
| Platelets, x10<sup>9</sup>/l * | 143 ± 52 | 110 ± 51 | <0.001 | 137 ± 54 | 102 ± 47 | <0.001 | 146 ± 52 | 116 ± 53 | <0.001 |
| AVB requiring PM implantation, n (%) |  |  |  |  |  |  |  |  |  |
| Among all patients | 60 (16.1) | 31 (33.7) | <0.001 | 60 (58.3) | 31 (77.5) | 0.035 | 0 | 0 | 1.000 |
| Among all patients without PM at baseline | 60 (18.2) | 31 (50.8) | <0.001 | 60 (100) | 31 (100) | 1.000 | 0 | 0 | 1.000 |
| New onset of LBBB after TAVI, n (%) |  |  |  |  |  |  |  |  |  |
| Among all patients | 60 (16.1) | 28 (30.4) | 0.002 | 1 (1.0) | 0 | 0.532 | 59 (21.9) | 28 (53.8) | <0.001 |
| Among patients without LBBB at baseline | 60 (16.6) | 28 (32.9) | <0.001 | 1 (1.0) | 0 | 0.537 | 59 (23.0) | 28 (60.9) | <0.001 |
| Length of hospital stay after TAVI, days † | 5 ± 2 | 6 ± 3 | <0.001 | 6 ± 4 | 7 ± 4 | 0.016 | 5 ± 2 | 6 ± 2 | <0.001 |
| **Echocardiographic characteristics at 30 days** |  |  |  |  |  |  |  |  |  |
| Mean gradient, mmHg * | 8 ± 4 | 14 ± 7 | <0.001 | 8 ± 3 | 13 ± 6 | <0.001 | 8 ± 5 | 14 ± 8 | <0.001 |
| Mean gradient ≥ 20, n (%) | 2 (0.6)<sup>#</sup> | 11 (13.1)<sup>#</sup> | <0.001 | 0<sup>#</sup> | 3 (8.6)<sup>#</sup> | 0.002 | 2 (0.8)<sup>#</sup> | 8 (16.3)<sup>#</sup> | <0.001 |
| Aortic regurgitation grade $\geq 3$, n (%) | 2 (0.6)<sup>#</sup> | 0<sup>#</sup> | 1.000 | 1 (1.1)<sup>#</sup> | 0<sup>#</sup> | 1.000 | 1 (0.4)<sup>#</sup> | 0<sup>#</sup> | 1.000 |
| **Echocardiographic characteristics at 1 year** |  |  |  |  |  |  |  |  |  |
| Mean gradient, mmHg * | 9 ± 3 | 12 ± 4 | <0.001 | 8 ± 3 | 11 ± 4 | 0.002 | 9 ± 3 | 12 ± 4 | <0.001 |
| Mean gradient $\geq 20$, n (%) | 1 (0.5)<sup>#</sup> | 3 (4.9)<sup>#</sup> | 0.040 | 0<sup>#</sup> | 1 (3.8)<sup>#</sup> | 0.321 | 1 (0.7)<sup>#</sup> | 2 (5.7)<sup>#</sup> | 0.094 |
| Aortic regurgitation grade $\geq 3$, n (%) | 1 (0.5)<sup>#</sup> | 0<sup>#</sup> | 1.000 | 0<sup>#</sup> | 0<sup>#</sup> | 1.000 | 1 (0.7)<sup>#</sup> | 0<sup>#</sup> | 1.000 |

\* Plus-minus values are mean ± SD; † Plus-minus values are median ± interquartile range; <sup>#</sup> Percentage is calculated from the available number of examinations performed. AVB, Atrioventricular block; LBBB, left bundle branch block; MEV, mechanically expandable valve; PM +, permanent pacemaker implanted; PM –, permanent pacemaker not implanted; SEV, self-expandable valve.

**Table 3.** Primary and secondary outcomes.

|  | SEV group<br>All<br>N = 373 | MEV group<br>All<br>N = 92 | *p*-value | SEV group<br>PM +<br>N = 103 | MEV group<br>PM +<br>N = 40 | *p*-value | SEV group<br>PM –<br>N = 270 | MEV group<br>PM –<br>N = 52 | *p*-value |
| --- | --- | --- | --- | --- | --- | --- | --- | --- | --- |
| Intraprocedural death, n (%) | 1 (0.3) | 0 | 1.000 | 0 | 0 | 1.000 | 1 (0.4) | 0 | 1.000 |
| Periprocedural myocardial infarction, n (%) | 3 (1.0)<sup>#</sup> | 2 (2.3)<sup>#</sup> | 0.310 | 1 (1.1)<sup>#</sup> | 0<sup>#</sup> | 1.000 | 2 (0.9)<sup>#</sup> | 2 (4.0)<sup>#</sup> | 0.167 |
| Myocardial injury not meeting MI criteria, n (%) | 178 (59.3)<sup>#</sup> | 70 (81.4)<sup>#</sup> | <0.001 | 62 (69.7)<sup>#</sup> | 30 (83.3)<sup>#</sup> | 0.178 | 116 (55.0)<sup>#</sup> | 40 (80.0)<sup>#</sup> | 0.001 |
| Overt CNS injury at 30 days, n (%) | 6 (1.6) | 2 (2.2) | 0.660 | 1 (1.0) | 1 (2.5) | 0.483 | 5 (1.9) | 1 (1.9) | 1.000 |
| Bleeding complications type 2- 4 at 30 days, n (%) | 13 (3.5) | 7 (7.6) | 0.081 | 3 (2.9) | 4 (10.0) | 0.096 | 10 (3.7) | 3 (5.8) | 0.448 |
| Acute kidney injury - stage 2 or 3, n (%) | 1 (0.3)<sup>#</sup> | 0<sup>#</sup> | 1.000 | 1 (1.0)<sup>#</sup> | 0 | 1.000 | 0<sup>#</sup> | 0<sup>#</sup> | 1.000 |
| Major vascular complications on TAVI day, n (%) | 3 (0.8) | 2 (2.2) | 0.258 | 2 (1.9) | 0 | 1.000 | 1 (0.4) | 2 (3.8) | 0.069 |
| Major vascular complications at 30 days, n (%) | 1 (0.3) | 2 (2.2) | 0.101 | 1 (1.0) | 0 | 1.000 | 0 | 2 (3.8) | 0.026 |
| Major non-vascular complications. on TAVI day, n (%) | 0 | 0 | 1.000 | 0 | 0 | 1.000 | 0 | 0 | 1.000 |
| Major non-vascular complications. at 30 days, n (%) | 0 | 0 | 1.000 | 0 | 0 | 1.000 | 0 | 0 | 1.000 |
| Major cardiac complications. on TAVI day, n (%) | 0 | 1 (1.1) | 0.198 | 0 | 1 (2.5) | 0.280 | 0 | 0 | 1.000 |
| Major cardiac complications. at 30 days, n (%) | 4 (1.1) | 0 | 1.000 | 1 (1.0) | 0 | 1.000 | 3 (1.1) | 0 | 1.000 |
| Incorrect positioning of a single prosthesis, n (%) | 6 (1.6) | 0 | 0.604 | 3 (2.9) | 0 | 0.096 | 3 (1.1) | 0 | 1.000 |
| Intervention related to the device or to a major vascular or access-related or cardiac structural complication, n(%) | 3 (0.8) | 1 (1.1) | 0.587 | 1 (1.0) | 1 (2.5) | 0.483 | 2 (0.7) | 0 | 1.000 |
| All-cause mortality at 30 days, n (%) | 12 (3.2) | 3 (3.3) | 1.000 | 3 (2.9) | 1 (2.5) | 1.000 | 9 (3.3) | 2 (3.8) | 0.694 |
| All-cause mortality at 1 year, n (%) | 50 (13.4) | 14 (15.2) | 0.773 | 20 (19.4) | 6 (15.0) | 0.452 | 30 (11.1) | 8 (15.4) | 0.445 |
| Cardiovascular mortality at 1 year, n (%) | 31 (8.3) | 11 (12.0) | 0.342 | 11 (10.7) | 4 (10.0) | 0.775 | 20 (7.4) | 7 (13.5) | 0.189 |
| All-cause mortality during follow-up, n (%) | 98 (26.3) | 46 (50.0) | 0.112 | 35 (34.0) | 19 (47.5) | 0.934 | 63 (23.3) | 27 (51.9) | 0.073 |
| Cardiovascular mortality during follow up, n (%) | 58 (15.5) | 31 (33.7) | 0.042 | 22 (21.4) | 12 (30.0) | 0.902 | 36 (13.3) | 19 (36.5) | 0.007 |
| Technical success, n (%) | 364 (97.6) | 89 (96.7) | 0.712 | 99 (96.1) | 39 (97.5) | 1.000 | 265 (98.1) | 50 (96.2) | 0.315 |
| Device success, n (%) | 344 (92.2) | 79 (85.9) | 0.057 | 95 (92.2) | 36 (90.0) | 0.666 | 249 (92.2) | 43 (82.7) | 0.030 |
| Early safety, n (%) | 288 (77.4) | 53 (57.6) | <0.001 | 39 (37.9) | 7 (17.5) | 0.019 | 249 (92.2) | 46 (88.5) | 0.370 |

### Primary outcomes

Overall, 144 all-cause deaths occurred over 1175 patient-years (mean follow-up ±SD = 2.5±1.7 years), which translates to 14.8 and 11.3 deaths per 100 patient-years in the MEV and SEV groups, respectively. The log-rank test showed higher mortality in the MEV group for all-cause deaths, but this difference did not reach statistical significance (*p=*0.112; RR: 1.345 [95%CI: 0.932-1.940]).

A total of 89 cardiovascular deaths occurred over follow-up, which translates to 10.0 MEV and 6.7 SEV deaths per 100 patient-years. The log-rank test showed significantly higher cardiovascular mortality in the MEV group (*p*=0.042; RR: 1.594 [95%CI: 1.013-2.508]) (Figure 1a).

**Figure 1a).**
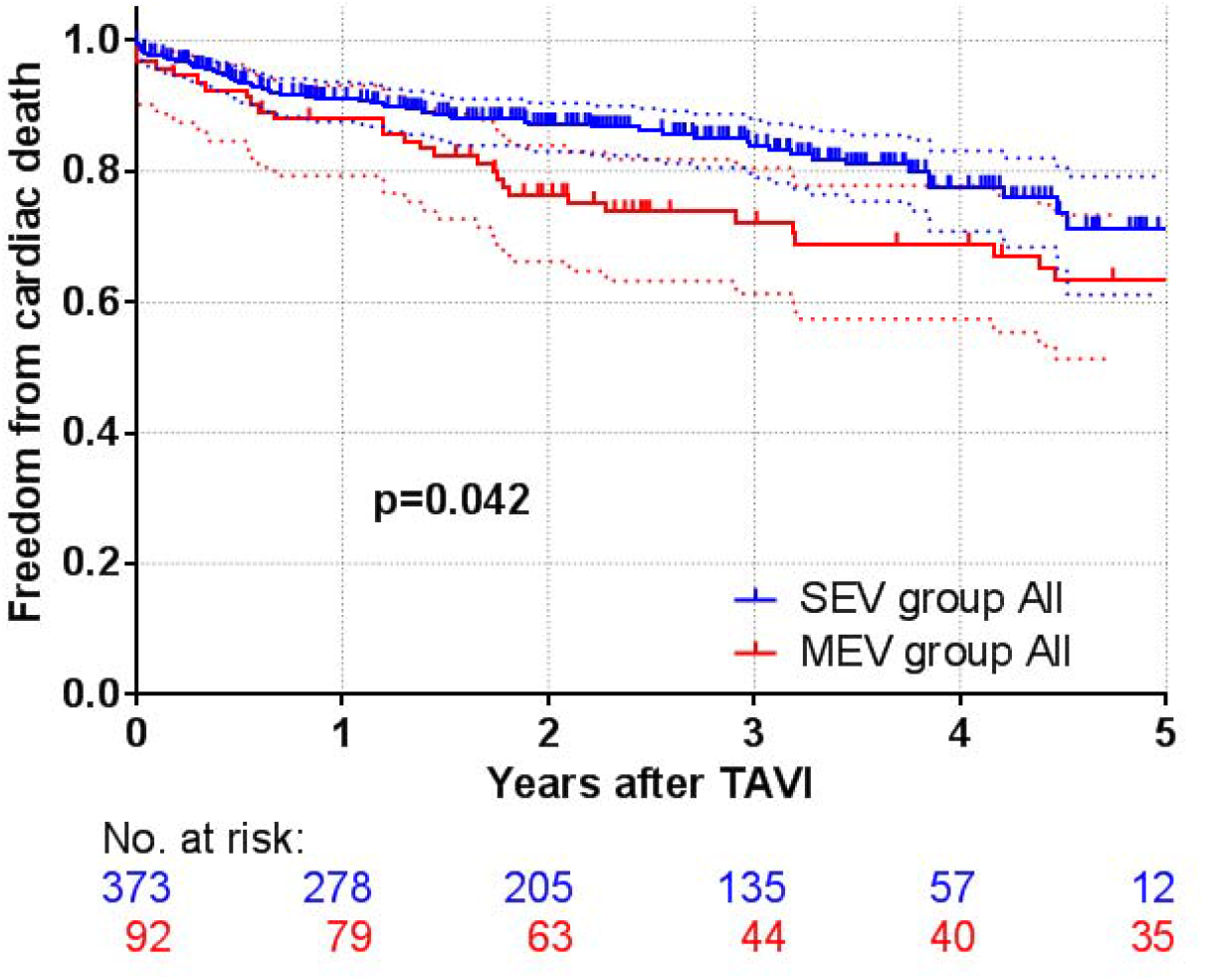
Cardiovascular mortality for self-expandable and mechanically expandable valves.

### Subanalysis of the studied patients

Overall, the Cox proportional hazards model identified the following factors as predictors of cardiovascular mortality: MEV implantation (*p*=0.021; RR: 1.71 [95%CI: 1.08-2.71]), male sex (*p*=0.003; RR: 1.93 [95%CI: 1.25-2.96]), higher STS score (*p*<0.001; RR: 1.05 [95%CI: 1.02-1.07]), and lower hemoglobin level before TAVI (*p*<0.001; RR: 0.98 [95%CI: 0.97-0.99]). Patients of the same age, with the same STS score and hemoglobin level before TAVI had a 1.7-fold higher risk of cardiovascular death with MEV than with SEV implants.

### Association of cardiovascular survival and pacemaker implantation after TAVI

We analyzed the survival of all patients according to the presence or absence of PPI, regardless of the implanted valve type. A total of 89 cardiovascular deaths occurred over follow-up, which translates to 9.4 and 6.8 deaths per 100 patient-years in the groups with and without PPI, respectively. The log-rank test did not reveal any mortality difference (*p*=0.140; RR: 1.38 [95%CI: 0.9-2.2]).

We performed further survival analysis comparing the MEV and SEV groups regarding the presence or absence of PPI. A total of 34 cardiovascular deaths occurred over follow-up in patients with any PPI (PPI before TAVI and new PPI after TAVI), which translates to 9.1 and 9.6 deaths per 100 patient-years in the MEV and SEV groups, respectively. A total of 24 cardiovascular deaths occurred over follow-up in patients with new PPI after TAVI, which translates to 9 and 10.3 deaths per 100 patient-years in the MEV and SEV groups, respectively. The log-rank test did not show any difference in cardiovascular mortality between groups (any PPI: *p*=0.908; RR: 0.961 [95%CI: 0.476-1.928]; new PPI after TAVI only: *p*=0.682; RR: 0.847 [95%CI: 0.372-1.889]) (Figure 1b).

**Figure 1b).**
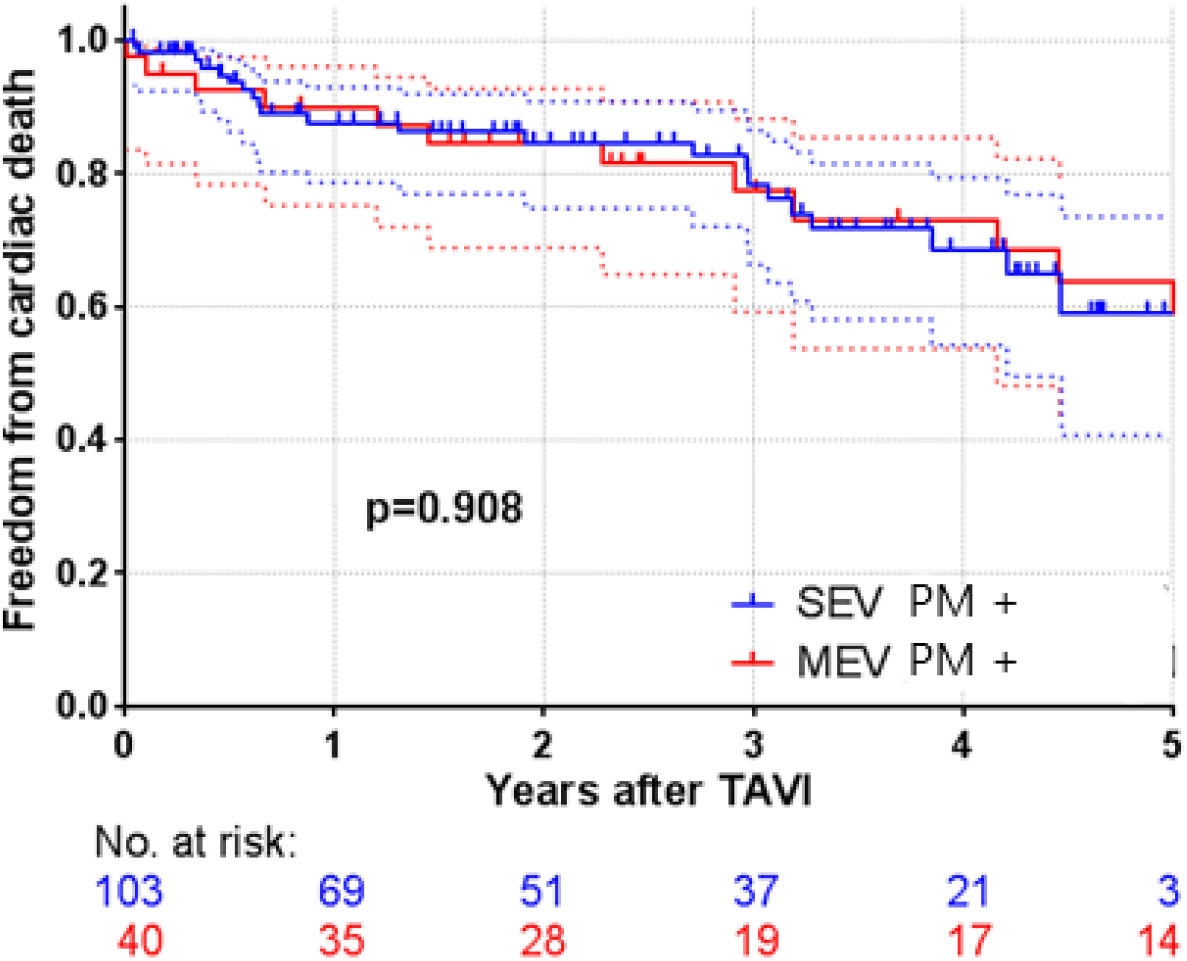
Cardiovascular mortality for self-expandable and mechanically expandable valves with permanent pacemaker implanted.

When we compared patients without PPI in the MEV and SEV groups, we found 55 cardiovascular deaths, which translates to 10.6 and 5.7 deaths per 100 patient-years in the MEV and SEV groups, respectively. The log-rank test showed significantly higher cardiovascular mortality in the MEV group (*p*=0.007; RR: 2.156 [95%CI: 1.213-3.831]) (Figure 1c).

**Figure 1c).**
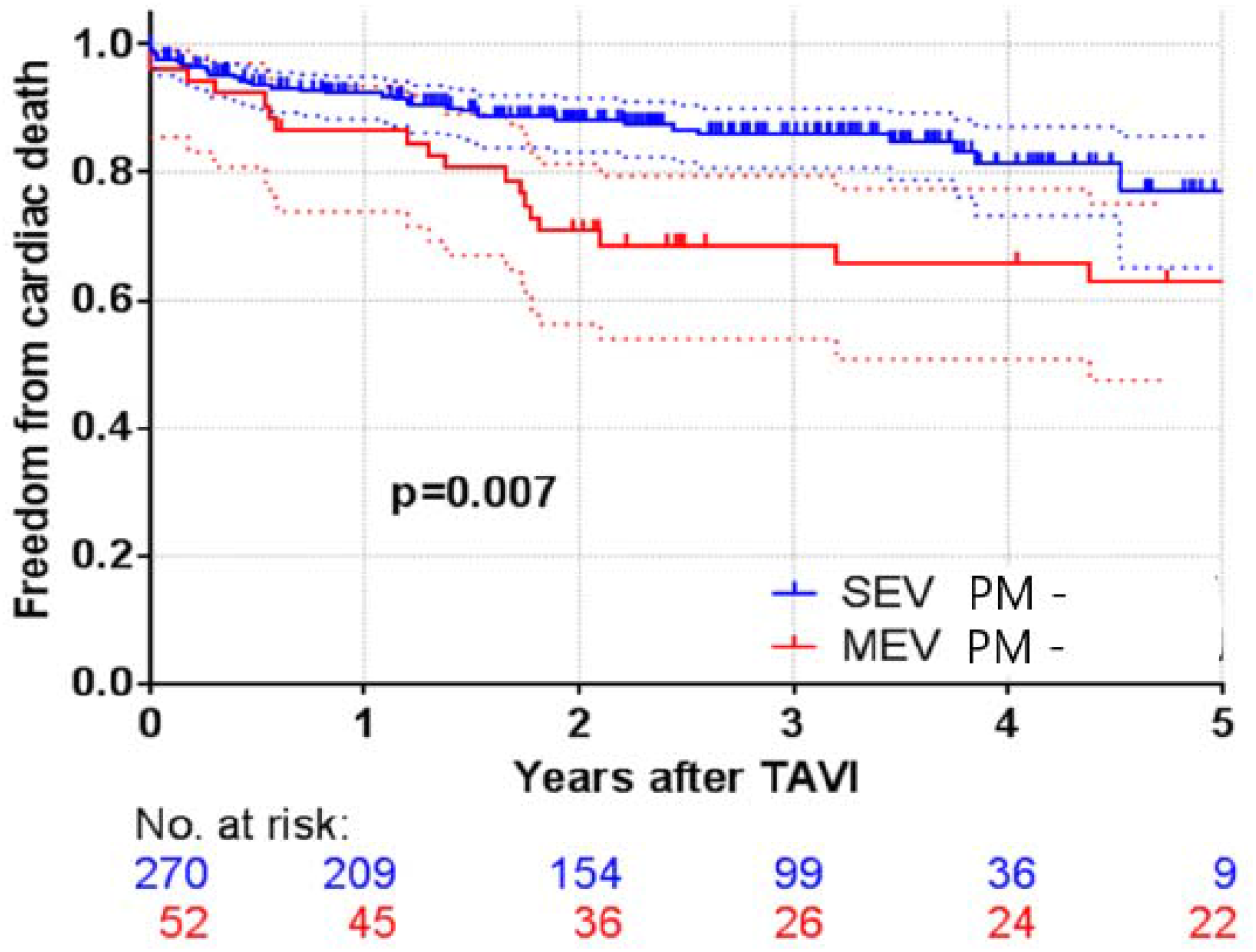
Cardiovascular mortality for self-expandable and mechanically expandable valves without permanent pacemaker implanted. SEV, self-expandable valve; MEV, mechanically expandable valve; PM +, permanent pacemaker implanted; PM -, permanent pacemaker not implanted.

### Subanalysis of patients without PPI

In patients without PPI, the Cox proportional hazards model identified the following predictors of cardiovascular mortality: MEV valve implantation (*p*=0.002; RR: 2.53 [95%CI: 1.41-4.53]), history of coronary artery disease (*p*=0.041; RR: 1.8 [95%CI: 1.02-3.15]), elevated STS score (*p*=0.012; RR: 1.04 [95%CI: 1.01-1.06]), and lower hemoglobin level before TAVI (*p*=0.002; RR: 0.97 [95%CI: 0.96-0.99]). Patients with the same history of coronary artery disease, STS score, and hemoglobin level before TAVI had a 2.5-fold higher risk of cardiovascular death with MEV than SEV implants.

## Discussion

Our single-center, retrospective, observational study reported two essential findings: (I) the use of MEV was independently associated with higher cardiovascular mortality than that of SEV; (II) MEV patients without PPI had a higher mortality risk than SEV patients without PPI. However, no mortality difference between the two groups was observed when a permanent pacemaker was implanted.

The mechanically expandable TAVI valve remains the only fully retrievable prosthesis with excellent results regarding minimal paravalvular leaking (10,15,16). Although the valve is no longer commercially available, thousands of prostheses have been implanted (6). Therefore, evaluation of the long-term outcomes of this valve is of utmost importance. Last year, the results of one observational study with a mean follow-up of 36 months (2) and those from a research correspondence with a median follow-up of 3.3 years (1) have been published. Both studies are consistent with the evidence for higher prevalence of valve endocarditis and thrombosis compared with other transcatheter valves. It is unclear whether these unfavorable outcomes also result in increased mortality over long-term follow-up.

Recently, a secondary analysis of the REPRISE III randomized clinical trial was presented. In contrast with our results, the findings of that analysis suggest that, at the 5-year follow-up, the Lotus® (i.e., MEV) valve had comparable outcomes to those of the CoreValve/Evolut R® (i.e., SEV)(6). In REPRISE III, 51.5% of SEV were prostheses of the first generation CoreValve, without the option to recapture and reposition, and in that regard unlike the second generation Evolut R. In the study by Giannini et al. (17), when compared with patients receiving the CoreValve, those treated with the Evolut R valve showed a significant survival benefit at one year (HR 1.80, 95% CI 1.01 to 3.23, *p*=0.046) and a significantly lower risk of PPI (22.3% and 35.0% for Evolut R and CoreValve, respectively; *p*=0.008). We believe that a significant proportion of the first generation prostheses in REPRISE III could have adversely affected the survival rate of the SEV group and thus, have mitigated the mortality difference we observed in our study.

Patients with NO-LBBB or requiring PPI after TAVI have a higher risk of death and rehospitalization for heart failure at one year (18) and over long-term follow-up than those without NO-LBBB or not receiving PPI (7,18). This has been primarily studied in balloon-expandable valves or SEV, where no significant survival difference according to the valve type was observed. Few studies included MEV in their analyses (7). In the largest randomized study (REPRISE III) (19) and the largest observational study (20) comparing MEV and SEV, the need for post-procedural PPI did not significantly impact survival within one year. In our study, we did not observe the negative prognostic association of MEV with PPI (*p*=0.648; RR: 0.832 [95%CI: 0.387-1.804]) or NO-LBBB (*p*=0.259; RR: 0.628 [95%CI: 0.262-1.430]). This suggests that the association of survival with post-MEV PPI may vary from that with PPI after other types of bioprosthesis.

The increased risk of all-cause death among patients receiving PPI can be related to cardiac and non-cardiac causes (7). Regarding these, mechanical injury to the LVOT that occurs during TAVI can play a role (21). In our study, the Lotus® valve caused an important mechanical injury to LVOT; this was documented by significantly higher number of new PPI (33.7% vs. 16.1% in the MEV and SEV groups, respectively; *p*<0.001) and NO-LBBB (30.4 vs. 16.1%, respectively; *p=*0.002). The long-lasting mechanical properties of MEV have been further supported by late-phase prosthesis expansion observed in a recent study by Kobari et al. (22).

Elevation of cardiac troponin level has been reported as a strong, independent predictor of 30-day mortality and a modest but significant predictor over 2 years of post-TAVI follow-up (23). We observed higher post-procedural troponin I levels in the MEV group than the SEV group, especially in the subgroups without PPI. In fact, the median troponin I levels even tended to be higher in the MEV group without PPI than in the SEV group with PPI (605 ± 1423 vs. 430 ± 1034 ng/l in the MEV and SEV groups, respectively; *p*=0.065).

The increased cardiac damage caused by the MEV, including higher incidence of new PPI, NO-LBBB, late-phase prosthesis expansion, and higher troponin I level after TAVI, raises the question regarding the impact of this long-term action of MEV on the LVOT in patients without PPI. Our results suggest that the survival of patients without PPI differs according to valve type. We assessed the relationship of new PPI and valve type using the Cox proportional hazards model. Despite the non-significant p-value (0.089), we found a 1.76-fold higher mortality risk for patients with SEV and new PPI compared to patients with the same valve without PPI. In contrast, we observed a 0.73-fold lower mortality risk for patients with MEV and new PPI compared to patients with the same valve without PPI. One possible explanation for the higher cardiovascular mortality in the MEV group without PPI may be late-onset conduction disturbances related to prolonged pressure of the prosthesis on the conduction system passing through the LVOT. Indeed, a study by Urena et al. suggested PPI as a protective factor against the occurrence of unexpected (sudden or unknown) death (24).

### Limitations

This study had several limitations. First, the retrospective, observational, single-center design has inherent limitations that should be considered before generalizing the results. Despite the non-randomized nature of our study, the groups were well balanced (Table 1). TAVI procedures (Table 3) were accompanied by a low rate of complications, without significant variation between the groups. The lower number of balloon valvuloplasties in the MEV group was possible because of valve properties (radial force and capability for full retrieval). The higher percentage of open surgical access stems from the preferred strategy during this period of our TAVI program, and it is most likely the cause of the lower hemoglobin levels after the procedure and longer hospital stays for patients with MEV (Table 2). A significantly higher composite endpoint of device success was driven by the higher frequency of the AVGm gradient > 20 mmHg in the MEV group. This higher post-procedural gradient with MEV has been observed in other studies (6,25) and is based on the intra-annular position of the prosthesis compared with the supra-annular placement of SEV. A significantly higher composite endpoint of early safety was driven by the higher frequency of new PPI associated with MEV. A high risk of conduction disturbances has also been associated with MEV (8,9). In our study, unlike MEV, AVGm> 20 mmHg and PPI were not identified as predictors of mortality.

Second, although we did not prove a difference in mortality between the MEV and SEV groups with implanted pacemakers, we cannot exclude the possibility of some variation. Nevertheless, our results indicated that the main mortality difference was observed in the groups without PPI.

Third, we did not find a difference of survival in the MEV group with and without PPI on the log-rank test; this would provide definitive proof of the association between PPI and survival.

## Conclusions

Despite several inherent limitations associated with the nonrandomized design of this study, our results suggest that a higher cardiovascular mortality rate was associated with mechanically expandable transcatheter valves over mid-term follow-up. On comparison of both groups according to the presence or absence of PPI, we observed a higher mortality risk in patients with MEV versus SEV without PPI. In contrast, we did not observe a mortality difference between the two groups when a permanent pacemaker was implanted.

## Data Availability

All data produced in the present work are contained in the manuscript

